# A Novel Linear B-spline Mixed Model for Repeated Measures (LB-MMRM) for Alzheimer’s Disease Clinical Study Data

**DOI:** 10.64898/2025.12.10.25341978

**Authors:** Kun Jin, William Chezem, Guoqiao Wang

**Affiliations:** Anavex Life Sciences Corp., New York, 10111, United States; Department of Neurology, Division of Biostatistics, Washington University in St. Louis, St Louis, 63130, United States

**Keywords:** MMRM, Linear B-Spline, Longitudinal data, Alzheimer’s disease, Unscheduled Visits

## Abstract

Mixed Models for Repeated Measures (MMRM) are widely used in neurological clinical trials, including Alzheimer’s disease studies, due to their robust statistical properties and regulatory acceptance. However, traditional MMRM treats time as a categorical variable, limiting its ability to incorporate unscheduled visits, harmonize trials with different visit schedules, or handle densely collected data from digital health technologies. To address these limitations, we propose a Linear B-Spline-based MMRM (LB-MMRM) model that uses time as a continuous variable while preserving compatibility with conventional MMRM when only scheduled visits are analyzed. LB-MMRM accommodates unscheduled visits by allocating observations to adjacent scheduled visits through spline-based weighting, thereby increasing effective sample size and statistical power. Simulation studies demonstrate that LB-MMRM maintains type I error control and improves power compared to traditional MMRM, particularly in scenarios involving irregular visit patterns or combined trial datasets. This approach offers a flexible and interpretable framework for modern clinical trials, supporting decentralized designs and integrated analyses without compromising regulatory standards.

## 1 Introduction

Mixed models for repeated measures (MMRM) are widely used as the primary analysis model in Alzheimer’s disease and other neurological clinical trials due to their robust statistical properties.^1-4^ A central aspect of this model is treating study visits as categorical variables, which imposes minimal constraints on the underlying statistical assumptions. The model provides estimates of treatment effect for each visit. The results of MMRM are easily interpretable for the regulatory decision, particularly for neurodegenerative disease, like Alzheimer’s since the treatment effect at the last visit is generally considered to be the primary result. However, this approach unfortunately limits its application in clinical trial data. Specifically, the MMRM cannot accommodate unscheduled visits, leading to a loss of information and increased missing data. Additionally, integrative studies cannot straightforwardly utilize MMRM for trials with different visit schedules. Lastly, as more wireless and densely spaced data (e.g., data collected by smartphones in burst design) become available, MMRM is impractical due to treating the time variable as categorical and creating hundreds of categories.^5^

To overcome these shortcomings, various models using time as a continuous variable have been proposed, including first-order time for linear mixed effects (LME) models and cubic time for (natural) cubic spline models, of both are parametric models.^4-7^ However, a major shortcoming of these methods is that their results are difficult to be interpreted in clinical trials. The LME model assumes linearity, which is generally not held for clinical trial data.^7-9^ The estimates of parameter in cubic spline model are not easy to interpret for the efficacy of the clinical trial data and can be inconsistent with MMRM estimates.^4, 10^

We aim to propose a hybrid approach, linear B-Spline-based MMRM (LB-MMRM) model, which uses time as a continuous variable. For the LB-MMRM with a 1-degree basis function, when only data from scheduled visits are used, it can yield the same results as traditional MMRM, thus providing a direct connection between the two approaches. We comprehensively evaluate the performance of LB-MMRM in comparison to MMRM for longitudinal data analysis. These models will be evaluated for several scenarios: (i) Unscheduled visits within scheduled visits (e.g., one month) are dropped in the MMRM model with categorical time. This scenario quantifies the disadvantage (i.e., loss of power) of using time as a categorical variable, which leads to more dropouts. (ii) Combining two clinical trials with different visit schedules (e.g., assessments every three months vs. every six months). This scenario quantifies the advantages of using time as a continuous variable to include more data in the analysis. The evaluation of model performance focuses on comparing type I error and power with the MMRM model.

The remainder of the paper presents the model formulations in Section 2, evaluates the behavior of each model through simulated hypothetical clinical trials in Section 3, and concludes with a discussion in Section 4.

## 2 Methods

### 2.1 Linear B-spline

B-splines are piecewise polynomial functions widely used for modeling smooth, flexible relationships in data.^11, 12^ Let *v*_*j*_, *j* = 0,1, …, *M*, represent the scheduled visits, *v*_0_ represents the baseline. The basis functions for linear B-spline (*B*_*j*_ (*t*)) can be generated as follows:

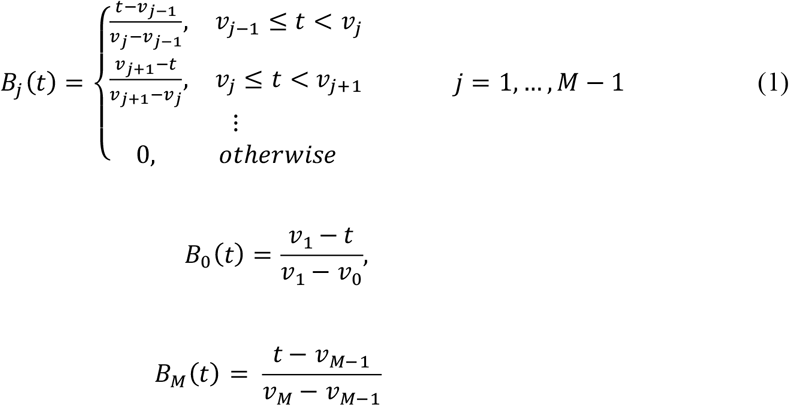

For the ease of demonstration, we use the following scheduled visits. [0,1,2,3,4]. *B*_*j*_ (*t*), *j* = 1,2,3,4 are selected to construct the LB-MMRM model, here *B*_*j*_ (*v*_*j*_ ) )=1, corresponding to scheduled visit *v*_*j*_. *B*_0_(*t*) is not selected due to the dependent variable is the change from baseline. Figure 1 illustrates the pattern of this “hat” shape of the B-spline basis function.

**Figure 1:**
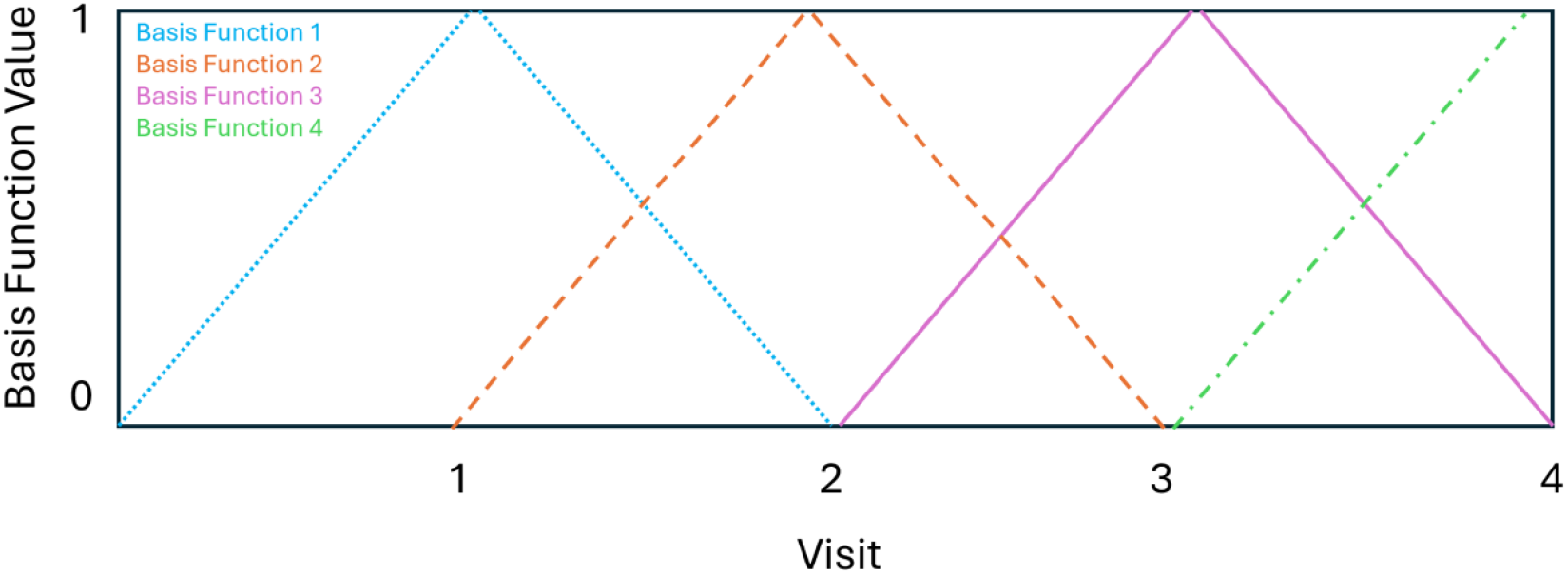
Illustration of the basis functions of B-spline. The first basis function peaks at the first knot (Visit 1) and the last basis function peaks at the last knot (Visit 4)

### 2.2 MMRM Model

Let, 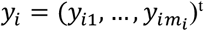, where 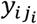 be *i* th patient change of assessment at visit *v*_*j*_ from baseline, *i* = 1, …, *n, m*_*i*_ ≤ *M*, where *n* is number of patients, *m*_*i*_ is the bumber of visits of *i*th patients. Let *X*_*i*_ be *m*_*i*_ × *M* desing matrix for fixed effect (e.g., *group, visit, group* × *visit*), *β*_*M*_ be vector of fixed effect parameters. Let 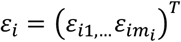 be the vector of the within-subject error and is assumed to follow 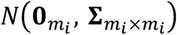, as is customarily in clinical trial analysis. Then the model for subject *i* is given by:

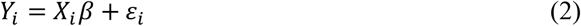

Here *X*_*i*_ = (x(*l*_*i*_, *l*_*j*_ )), *l*_*i*_ = 1, …, *m*_*i*_, *l*_*j*_ = 1, …, *M*, can be descripted is the following

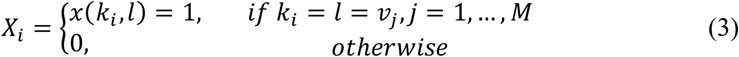

Denote 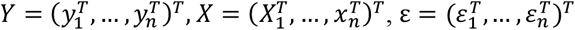

The MMRM model is

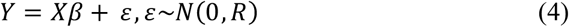

Where *Y*, ε are 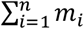 vectors, and *X* is the 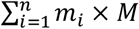 matrix.

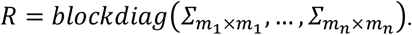

For the testing of the primary efficacy, let’s assume two groups, *D* as drug group and *P* as placebo for simplicity. Let 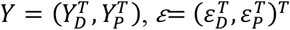, then the MMRM model is

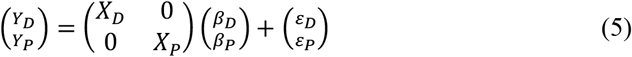

Where *β*_*D*_, *β*_*P*_ are parameters of the fixed effect, and 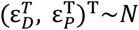(0, *blockdiag*(*R*_*D*_, *R*_*P*_)). The REML or ML methods are applied to the log likelihood function of 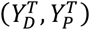 to get estimates of the fixed effect, 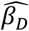 and 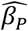 and the variance-covariance matrices, 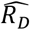 and 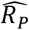.

### 2.3 Linear B-spline MMRM

An important feature of both REML and ML algorithms for estimating the MMRM parameters is that they require only the design matrices *X*_*D*_ and *X*_*P*_, each with *M* columns corresponding to the number of visits *M*. The number of rows, that corresponds to the length of vector *Y*, can be as many as observed.

We will use *B*_*D*_(*t*), and *B*_*p*_(*t*), (*j* = 1, …, *M*) in 2.1 to construct new design matrices, *B*_*D*_ (*t*) = (*B*_*D*,1_, …, *B*_*D,M*_) and *B*_*p*_(*t*) = (*B*_*p*,1_, …, *B*_*p,M*_) to replace *X*_*D*_ and *X*_*P*_ in (5) to accommodate the continuous visits, *t*.

In the following discussion, we will not distinguish between *B*_*D*_ and *B*_*P*_ for simplicity. We will only refer to the general case and *B*(*t*), and the discussion applies to both *B*_*D*_ and *B*_*P*_ .

Let 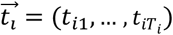 be a vector of continuous time in the study interval [*v*_0_, *v*_*M*_]. Some of 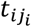 are visit *v*_*l*_,(*l* = 1, …, *M*), others are unscheduled visits in between visits. Let 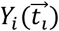 denote the vector of changes from baseline at *t* for the *i*th patient.

For this patient, the corresponding design matrix is 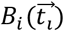, the matrix of base functions evaluated at 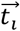.

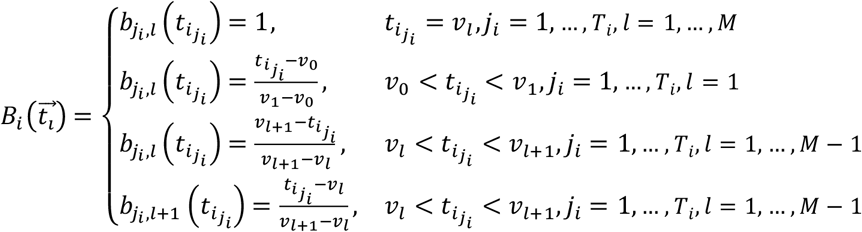

If there are no unscheduled visits, 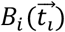 will be the same as *X*_*i*_ in the formula (3). When unscheduled visits occur, 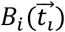 will split the unscheduled visit data to adjacent scheduled visits.

- If an unscheduled visit falls between baseline (zero) and the first scheduled visit, *B* will allocate the observed data to the column *v*_*l*_ with a weight:

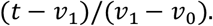
- If an unscheduled visit occurs between *v*_*l*_ and *v*_*l*+1_ (where *l* = 1, …, *M* ™ 1), the observed data will be split between the data to column of *v*_*l*_ and *v*_*l*+1_ with weights:

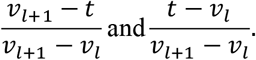

By anchoring visits as the basis for the splines, the matrix 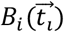 will have *M* columns aligned with the commonly used variance–covariance matrix structure.

When longitudinal data without unscheduled visits are used, this model produces the same results as the traditional MMRM method. This consistency is essential for regulatory actions. When unscheduled visits are present, the new model gains more power by incorporating additional data while maintaining similar statistical inference and generating comparable results for regulatory purposes.

### 2.4 Simulation Studies

To assess the Type I error and power of the LB-MMRM model, we performed 5,000 Monte Carlo replications under a two-arm design with 150 patients per group. Data were generated using the R functions **SPR::sim_dat** and **SPR::dropout**, assuming an AR(1) covariance structure with variance = 1, correlation = 0.8, and a true treatment effect (Δ) of 0.3 for power scenarios. Missing data were introduced under both MAR and MNAR mechanisms at rates of 10%, 20%, and 30%. Additionally, unscheduled visits were incorporated, with MAR scenarios including 1 or 2 extra visits per interval and MNAR scenarios including 2 or 3. The proportion of patients with unscheduled visits varied across 50%, 80%, and 100%. These design factors allowed us to evaluate the robustness of LB-MMRM under realistic clinical trial conditions with varying dropout patterns and visit irregularities.

## 3 Results

Under the MAR assumption for missing data, with 50% of patients having unscheduled observations between each visit interval the LB-MMRM model consistently outperforms the standard MMRM model in terms of statistical power regardless of the amount of missing data, with the difference between models increasing as the amount of missing data increases. At 0% missing data and 1 unscheduled visit per subject, the LB-MMRM’s power at Visit 4 is 0.746, compared to 0.741 for the MMRM model (Table 1), with comparable Type I error rates of 0.054 for the LB-MMRM model and 0.052 for the MMRM model. The difference between the two models increases as the amount of missing data at scheduled visits increases: at 30% missing data and two unscheduled visits per subject the LB-MMRM’s statistical power is 0.670 compared to 0.553 in the MMRM model (Table 1), while the Type I error rate remains comparable between the two models: 0.044 for the LB-MMRM model versus 0.046 for the MMRM model. Similarly, the difference in power between the two models also increases as the number of unscheduled observations between each interval increases (Table 2).

**Table 1.** Power and Type I error comparison for missing at random, 50% Patients with unscheduled observations of 1 or 2 within each interval.

| Missing | Model | Statistic | Visit |  |  |  |  |  |  |  |
| --- | --- | --- | --- | --- | --- | --- | --- | --- | --- | --- |
|  |  |  | V1 |  | V2 |  | V3 |  | V4 |  |
|  |  |  | 1 un | 2 un | 1 un | 2 un | 1 un | 2 un | 1 un | 2 un |
| 0% | LB-MMRM | Power | 0.624 | 0.66 | 0.666 | 0.697 | 0.702 | 0.729 | 0.746 | 0.761 |
|  |  | Type I Error | 0.051 | 0.048 | 0.043 | 0.05 | 0.051 | 0.048 | 0.054 | 0.05 |
|  | MMRM | Power | 0.606 | 0.614 | 0.653 | 0.672 | 0.695 | 0.703 | 0.741 | 0.746 |
|  |  | Type I Error | 0.05 | 0.051 | 0.046 | 0.048 | 0.052 | 0.047 | 0.052 | 0.049 |
| 10% | LB-MMRM | Power | 0.614 | 0.632 | 0.648 | 0.7 | 0.679 | 0.719 | 0.723 | 0.739 |
|  |  | Type I Error | 0.055 | 0.047 | 0.05 | 0.049 | 0.052 | 0.048 | 0.05 | 0.045 |
|  | MMRM | Power | 0.567 | 0.539 | 0.598 | 0.642 | 0.625 | 0.653 | 0.672 | 0.691 |
|  |  | Type I Error | 0.053 | 0.045 | 0.05 | 0.051 | 0.051 | 0.049 | 0.048 | 0.047 |
| 20% | LB-MMRM | Power | 0.594 | 0.599 | 0.611 | 0.691 | 0.632 | 0.694 | 0.649 | 0.713 |
|  |  | Type I Error | 0.056 | 0.048 | 0.051 | 0.052 | 0.051 | 0.049 | 0.053 | 0.045 |
|  | MMRM | Power | 0.517 | 0.453 | 0.53 | 0.619 | 0.543 | 0.611 | 0.554 | 0.631 |
|  |  | Type I Error | 0.052 | 0.046 | 0.05 | 0.053 | 0.051 | 0.048 | 0.051 | 0.046 |
| 30% | LB-MMRM | Power | 0.585 | 0.542 | 0.585 | 0.674 | 0.578 | 0.67 | 0.563 | 0.67 |
|  |  | Type I Error | 0.053 | 0.045 | 0.05 | 0.05 | 0.052 | 0.044 | 0.051 | 0.044 |
|  | MMRM | Power | 0.472 | 0.362 | 0.464 | 0.594 | 0.46 | 0.556 | 0.455 | 0.553 |
|  |  | Type I Error | 0.05 | 0.044 | 0.047 | 0.052 | 0.046 | 0.044 | 0.048 | 0.046 |

**Table 2.** Power and Type I error comparison for missing at random, 100% Patients with unscheduled observations of 1 or 2 within each interval.

| Missing | Model | Statistic | Visit |  |  |  |  |  |  |  |
| --- | --- | --- | --- | --- | --- | --- | --- | --- | --- | --- |
|  |  |  | V1 |  | V2 |  | V3 |  | V4 |  |
|  |  |  | 1 un | 2 un | 1 un | 2 un | 1 un | 2 un | 1 un | 2 un |
| 0% | LB-MMRM | Power | 0.637 | 0.662 | 0.669 | 0.705 | 0.722 | 0.742 | 0.751 | 0.760 |
|  |  | Type I Error | 0.049 | 0.049 | 0.051 | 0.049 | 0.047 | 0.046 | 0.047 | 0.049 |
|  | MMRM | Power | 0.600 | 0.613 | 0.647 | 0.657 | 0.693 | 0.702 | 0.735 | 0.734 |
|  |  | Type I Error | 0.047 | 0.052 | 0.051 | 0.053 | 0.050 | 0.049 | 0.046 | 0.050 |
| 10% | LB-MMRM | Power | 0.657 | 0.670 | 0.684 | 0.715 | 0.724 | 0.749 | 0.747 | 0.766 |
|  |  | Type I Error | 0.045 | 0.045 | 0.053 | 0.050 | 0.049 | 0.052 | 0.049 | 0.050 |
|  | MMRM | Power | 0.563 | 0.530 | 0.582 | 0.633 | 0.628 | 0.652 | 0.661 | 0.680 |
|  |  | Type I Error | 0.044 | 0.047 | 0.049 | 0.056 | 0.047 | 0.053 | 0.047 | 0.050 |
| 20% | LB-MMRM | Power | 0.670 | 0.663 | 0.674 | 0.719 | 0.709 | 0.750 | 0.714 | 0.758 |
|  |  | Type I Error | 0.046 | 0.049 | 0.048 | 0.048 | 0.045 | 0.053 | 0.051 | 0.049 |
|  | MMRM | Power | 0.517 | 0.447 | 0.528 | 0.606 | 0.552 | 0.599 | 0.556 | 0.607 |
|  |  | Type I Error | 0.050 | 0.046 | 0.049 | 0.057 | 0.043 | 0.049 | 0.046 | 0.050 |
| 30% | LB-MMRM | Power | 0.675 | 0.647 | 0.670 | 0.728 | 0.681 | 0.742 | 0.662 | 0.743 |
|  |  | Type I Error | 0.044 | 0.048 | 0.047 | 0.047 | 0.046 | 0.053 | 0.047 | 0.046 |
|  | MMRM | Power | 0.484 | 0.354 | 0.478 | 0.576 | 0.476 | 0.547 | 0.447 | 0.536 |
|  |  | Type I Error | 0.046 | 0.048 | 0.050 | 0.058 | 0.042 | 0.047 | 0.044 | 0.049 |

The same pattern continues for the results under the missing not at random assumption. At 80% of patients with unscheduled observations, 0% missing data, and two unscheduled observations per interval, the statistical power of the LB-MMRM model at the first visit is 0.662 compared to 0.613 for the MMRM model (Table 3). This difference increases as the amount of missing data increases and at 30% missing data and at the last visit, the power of the LB-MMRM model is 0.661 versus 0.439 for the MMRM model, while Type I error (0.045) is also lower than in the corresponding MMRM model (0.049; Table 3).

**Table 3.** Power and Type I error comparison for missing not at random, 80% Patients with unscheduled observations of 2 or 3 within each interval.

| Missing | Model | Statistic | Visit |  |  |  |  |  |  |  |
| --- | --- | --- | --- | --- | --- | --- | --- | --- | --- | --- |
|  |  |  | V1 |  | V2 |  | V3 |  | V4 |  |
|  |  |  | 2 un | 3 un | 2 un | 3 un | 2 un | 3 un | 2 un | 3 un |
| 0% | LB-MMRM | Power | 0.662 | 0.672 | 0.705 | 0.710 | 0.742 | 0.746 | 0.760 | 0.775 |
|  |  | Type I Error | 0.049 | 0.050 | 0.049 | 0.051 | 0.046 | 0.049 | 0.049 | 0.053 |
|  | MMRM | Power | 0.613 | 0.619 | 0.657 | 0.659 | 0.702 | 0.703 | 0.734 | 0.740 |
|  |  | Type I Error | 0.052 | 0.046 | 0.053 | 0.050 | 0.049 | 0.049 | 0.050 | 0.048 |
| 10% | LB-MMRM | Power | 0.591 | 0.635 | 0.671 | 0.682 | 0.687 | 0.727 | 0.706 | 0.743 |
|  |  | Type I Error | 0.045 | 0.049 | 0.057 | 0.053 | 0.050 | 0.054 | 0.049 | 0.050 |
|  | MMRM | Power | 0.478 | 0.483 | 0.595 | 0.523 | 0.601 | 0.623 | 0.624 | 0.643 |
|  |  | Type I Error | 0.047 | 0.046 | 0.061 | 0.049 | 0.050 | 0.059 | 0.047 | 0.050 |
| 20% | LB-MMRM | Power | 0.526 | 0.577 | 0.655 | 0.652 | 0.638 | 0.706 | 0.645 | 0.704 |
|  |  | Type I Error | 0.043 | 0.045 | 0.062 | 0.054 | 0.047 | 0.053 | 0.042 | 0.048 |
|  | MMRM | Power | 0.346 | 0.352 | 0.550 | 0.370 | 0.524 | 0.560 | 0.520 | 0.547 |
|  |  | Type I Error | 0.042 | 0.041 | 0.062 | 0.041 | 0.046 | 0.061 | 0.041 | 0.051 |
| 30% | LB-MMRM | Power | 0.463 | 0.525 | 0.637 | 0.625 | 0.589 | 0.669 | 0.581 | 0.661 |
|  |  | Type I Error | 0.042 | 0.041 | 0.066 | 0.057 | 0.046 | 0.054 | 0.044 | 0.045 |
|  | MMRM | Power | 0.229 | 0.248 | 0.522 | 0.224 | 0.452 | 0.503 | 0.413 | 0.439 |
|  |  | Type I Error | 0.037 | 0.044 | 0.066 | 0.035 | 0.044 | 0.061 | 0.038 | 0.049 |

## 4 Discussion and Conclusion

The proposed LB-MMRM model offers a natural extension of the traditional Mixed Model for Repeated Measures (MMRM) while introducing key innovations that enhance its applicability in modern clinical trial settings. By using continuous time and maintaining identical outputs when scheduled visit data are analyzed, LB-MMRM ensures compatibility with established methodologies, facilitating adoption without disrupting current practices.

A major advantage of LB-MMRM is its ability to incorporate unscheduled visits, which increases the effective sample size and improves statistical power. This feature is particularly relevant for trials where patient adherence varies, or additional data points become available outside the predefined schedule. Furthermore, the model supports the inclusion of home-based measurements, provided appropriate calibration is applied. This capability aligns with the growing trend toward decentralized trials and remote monitoring, enabling richer datasets and more patient-centric designs.

LB-MMRM also provides a robust framework for comparing studies or treatment arms with differing visit schedules, such as integrated summaries of efficacy or analyses involving external historical controls. This flexibility addresses a long-standing challenge in harmonizing data across heterogeneous sources and strengthens the utility of real-world evidence in regulatory and research contexts.

In conclusion, LB-MMRM combines methodological rigor with practical flexibility, offering a powerful tool for longitudinal data analysis. Future work should focus on optimizing computational efficiency, validating calibration procedures for home-based devices, and exploring sensitivity analyses for missing data mechanisms. These advancements will further enhance support for innovative trial designs and improve the robustness of efficacy evaluations.

## Data Availability

All data produced in the present work are contained in the manuscript

